# PM_2.5_ components mixture and atherosclerotic cardiovascular disease mortality: a national analysis of Medicare enrollees

**DOI:** 10.1101/2024.03.23.24304739

**Authors:** Tszshan Ma, Pablo Knobel, Michael Hadley, Elena Colicino, Heresh Amini, Alex Federman, Joel Schwartz, Kyle Steenland, Maayan Yitshak Sade

**Author notes:** Tszshan Ma and Pablo Knobel contributed equally to this paper. **Corresponding author:** Maayan Yitshak Sade Department of Environmental Medicine and Public Health Icahn School of Medicine at Mount Sinai New York, New York, United States.

## Abstract

Fine particulate matter (PM_2.5_) exposure is adversely linked to atherosclerotic cardiovascular disease (ASCVD). However, most studies focused on PM_2.5_ mass rather than its chemical composition. PM_2.5_’s individual chemical components can have distinct, cumulative, and potentially synergistic health impacts. We investigated the associations of PM_2.5_’s composition and sources with ASCVD mortality, considering the combined associations and regional variations in the US. We used data from the Centers for Medicare and Medicaid Services, (65,838,403 person-years) from 2000 to 2016. We estimated PM_2.5_ exposure using machine-learning models and attributed components to five source categories. We used Poisson survival models to assess the associations with the source categories. Higher ASCVD mortality risk (RR [95% CI] per interquartile range increase) was associated with oil combustion (1.050[1.049;1.051]), industrial (1.054[1.052;1.056]), coal/biomass burning (1.064[1.062;1.067]), and traffic sources (1.044[1.042;1.046]). Comparing source-specific effects within each region, oil combustion effects were more pronounced in the East and Midwest, and coal/biomass burning effects were more pronounced in the West and Southwest. In conclusion, we found higher ASCVD mortality risk associated with PM_2.5_, with differential effects across sources and US regions. These associations persisted even after limiting our sample to ZIP code-years with PM_2.5_ <9 μg/m^3^ - the National Ambient Air Quality Standards (NAAQS). This highlights the importance of consideration of local population characteristics and exposure patterns when assessing health risks associated with PM_2.5_.

## 1. Introduction

Research has shown that lifestyle factors such as physical activity, weight loss, smoking, diet, and stress^1^ and environmental factors, such as greenness, temperature, and air pollution^2^, play a critical role in developing cardiovascular disease (CVD). Particulate matter in the atmosphere with an aerodynamic diameter less than or equal to 2.5 μm (PM_2.5_) has been conclusively linked to the development of cardiovascular diseases^3^, particularly Atherosclerotic cardiovascular disease (ASCVD)^4^. ASCVD encompasses two primary conditions: ischemic heart disease (IHD) and cerebrovascular disease (predominantly ischemic stroke), which are the top and third leading causes of death globally^5^. In 2019, PM_2.5_ was responsible for 2.48 million CVD deaths and 60.91 million CVD-related disability-adjusted life years (DALYs) globally ^6^. By setting limits on PM_2.5_ concentrations, the National Ambient Air Quality Standards (NAAQS) aim to mitigate the health risks of PM_2.5_ exposure among the population. However, despite the protective intent of the NAAQS, the standards for PM_2.5_ (<9 μg/m^3^) are still higher than the recommendations set by the World Health Organization (WHO) (<5 μg/m^3^). The WHO’s guidelines are more stringent, reflecting the latest research linking lower levels of PM2.5 exposure to significant health benefits. The disparity between the NAAQS and WHO guidelines highlights a critical area for policy improvement. Lowering the NAAQS for PM_2.5_ to align more closely with WHO recommendations could potentially reduce the burden of cardiovascular diseases further, showcasing the ongoing need to adjust environmental standards based on evolving scientific evidence.

Air pollution results from combustion and complex chemical reactions involving various emissions. Although research often focuses on the mass of PM_2.5_, its chemical composition is crucial. The composition of PM_2.5_ is complex, and the physical structure, chemical composition, and source of its components are diverse^7^. Therefore, PM_2.5_’s individual chemical components can have distinct, cumulative, and potentially synergistic health impacts. A few studies have documented the links between some components of PM_2.5_ and various cardiovascular health-related outcomes. For example, elemental or black carbon, primarily derived from combustion processes, has been linked to increased hospitalizations and deaths due to cardiovascular issues^8^. Potassium, a marker of biomass burning, has been linked with cardiovascular and respiratory health risks^9^. Vanadium, nickel, and particles from oil combustion have been associated with mortality from cardiovascular diseases^10^. A mixture analysis revealed potassium, organic carbon, and iron had the strongest associations with overall, cardiovascular, and respiratory mortality ^11^. Another mixture analysis reported an association with all-cause and cardiovascular mortality, with the largest weights for organic carbon, nickel, zinc, sulfate, and vanadium^12^. However, these results are often inconsistent^13^, and evidence for the combined effects of components is very limited^14^.

In the United States, the distribution of PM_2.5_ and its chemical components is heterogeneous, revealing a pattern of exposure that varies from one community to another^15^. This variability in PM_2.5_ exposure has implications as studies have consistently demonstrated that specific subgroups within the population, such as racial/ethnic minorities, the elderly population, and individuals with low socioeconomic status (SES), are disproportionately affected by adverse cardiovascular disease (CVD) outcomes^16^. Differences amongst communities can also be present due to differences in customs, traditions, and lifestyle - often a combination of personal sociodemographic identities and geographic context.

Investigating the combined effects of PM_2.5_ chemical components poses two major challenges: first, the requirement of component-specific high-quality data - with high resolution and low uncertainty - to assign exposures with minimal misclassification. Additionally, high-quality exposure data must be available over large areas to allow for comparisons amongst communities. Second, the analysis of this data requires statistical methods specifically designed for handling the highly correlated structure of component data and extracting meaningful estimates. This study leverages state-of-the-art PM_2.5_ and components spatial datasets with high resolution and low uncertainly as well as statistical methods capable of handling the complexities of the data to examine the relationships between PM_2.5_ and its components and ASCVD mortality, considering their combined effects, as well as the variations among regions across the United States. To this end, we used factorization of components into sources to link specific sources and combinations of components with ASCVD mortality.

## 2. Methods

### 2.1. Study Population and Mortality Data

We obtained two nationwide databases from the Centers for Medicare and Medicaid Services (CMS) for the period 2000-2016, including the Master Beneficiary Summary File (MBSF) and the Medicare National Death Index (NDI) segment. Medicare is the largest health insurance provider in the US, covering over 95% of the population aged 65 years and above. This open cohort enrolls new members annually and covers most older adults in the US. The MBSF contains enrollment records for each Medicare beneficiary, including age, sex, race, Medicaid eligibility, death date (if any), and ZIP residence code, and is updated annually. The NDI database is a centralized database of death records in state vital statistics offices. The NDI includes information about the location of death (state), the date of death, a single underlying cause of death, and up to twenty additional multiple causes for each death. The outcome tested in this study (extracted from the single underlying cause of death) was atherosclerotic cardiovascular disease (ASCVD) mortality, defined as either ischemic heart disease or cerebral infarction mortality (ICD-10 codes I20, I21, I22, I23, I24, I25, I63, I65, I66). Study subjects entered the cohort on January 1^st^ of the year after they enrolled in Medicare and were followed for each calendar year until death, end of enrollment in Medicare, or end of follow-up, whichever came first.

The Center for Medicare and Medicaid Services (CMS) approved this study under the data use agreement (#RSCH-2020-55,733), the Institutional Review Board of Emory University (#STUDY00000316), and the Institutional Review Board of Mount Sinai (STUDY 20–01344). Additionally, a waiver of informed consent was granted. The Medicare dataset was stored and analyzed in the Rollins High-Performance Computing (HPC) Cluster at Emory University in compliance with the Health Insurance Portability and Accountability Act (HIPAA).

### 2.2 Exposure Data

We obtained annual mean predictions for 15 PM_2.5_ components, including sulfate (SO_4_^2-^), nitrate (NO_3_^-^), ammonium (NH_4_^+^), organic carbon (OC), elemental carbon (EC), zinc (Zn), vanadium (V), potassium (K), silicon (Si), lead (Pb), nickel (Ni), iron (Fe), copper (Cu), calcium (Ca), and bromine (Br), at 50-m spatial resolution for urban areas and 1-km resolution for non-urban areas across the contiguous US (2000-2016) from super-learning approach models (i.e. ensemble of ensemble learning)^17,18^. The cross-validated R^2^ values for individual components ranged from 0.821 (Br) to 0.975 (SO_4_^2-^). Details about the PM_2.5_ components’ exposure data can be found elsewhere^17,18^, and the data are publicly accessible via the NASA Socioeconomic Data and Applications Center (NASA SEDAC) website ^19,20^. We calculated population-weighted annual averages for each pollutant for each ZIP code (i.e., the finest spatial resolution in Medicare data), considering that the population is not distributed evenly within the ZIP code. Specifically, we re-gridded the modeled exposure dataset at 1 km resolution into a common grid consistent with the finest resolution (30 arc-seconds, ∼ 1 km at the equator) of the population density dataset from NASA’s Gridded Population of the World. The exposure data within each ZIP code polygon was aggregated by population-weighted average for each year over the study period (2000-2016) and assigned to each Medicare beneficiary based on the calendar year and ZIP code of residences that are tracked and updated annually.

### 2.3 Covariates

We obtained individual-level demographics (age, sex, and race) and Medicaid insurance status from the Medicare MBSF. We created a 5-year age-at-entry group variable based on individual age at entry into the Medicare cohort. To further adjust our models we obtained ZIP code-level socioeconomic characteristics (population density, % Black population, % Black Hispanic population, % population aged 65 or above living below the poverty line, % population receiving public assistance, % population living in rental house or apartment, median household income, and percentage population with less than a high school education), meteorological variables (annual mean summer and winter temperatures), land-use variables (normalized difference vegetation index (NDVI) values), county-level health care capacity indicators (number of hospitals and active medical doctors per 1,000 people), and a geographic region variable categorized as five US regions (West, Midwest, Northeast, Southeast, and Southwest, as shown in **Supplementary figure S1**). Details and data sources of the covariates are described in Ma et al. (2022)^21^.

### 2.4 Statistical Methods

#### Descriptive analysis

Descriptive statistics were used for all included variables for the whole US and stratified for each US region. Pearson correlation was used to examine the correlations between PM_2.5_ components.

#### Non-negative matrix factorization (NMF) of 15 PM_2.5_ components

To evaluate the independent associations of PM_2.5_ sources, we used NMF to group the components into source categories. This method aims to identify PM_2.5_ source categories containing a mixture of pollutants associated with higher ASCVD mortality risks. Due to its non-negative constraint, NMF is an efficient method for dimension reduction of a matrix of PM_2.5_ chemical components into a lower dimension non-negative matrix that best approximates the original components data ^15,22^. We tested for possible 4 to 6 factors and conducted 100 multiple runs to obtain the best factorization fit. We selected five factors based on the inflection point on the residuals sum of squares curve, a visual inspection of the mixture coefficient matrix, and an assessment of the correlation between the factors. We then labeled the source categories based on the high-loading components known as tracers of PM_2.5_ sources.

#### Associations of PM_2.5_ sources and ASCVD mortality

We used Poisson survival analyses ^23^ using the Anderson– Gill formulation with adjusted individual-level covariates (sex, race, Medicaid eligibility, 5-year categories of age at study entry), as well as the area-level covariates (socioeconomic, meteorological, land use, and health care use variables). We generated an artificial Poisson observation for each subject in the risk set at each non-censored year with an outcome of zero for all person-years except those experiencing the event in that calendar year. Person-years (with the same sex, race, Medicaid eligibility, and 5-year age categories at study entry) in the same ZIP code and calendar year were aggregated and treated as interchangeable in the analysis. We allowed a piecewise constant hazard for each year using indicator variables for each calendar year. Finally, we considered the derived 5 NMF loading factors as co-exposures and fitted the outcome model as a Poisson regression, adjusting for the same covariates described. We then repeated the models within each US region separately, to identify potential differential effects by geographical region (West, Midwest, Northeast, Southeast, and Southwest) and overall. Results are presented in RR with 95% CI per IQR increase for the NMF factor.

#### Focusing on zip codes-years below 9 μg/m^3^ PM_2.5_

We subseted our dataset to zip code-years below 9 μg/m^3^ PM_2.5_ to evaluate the associations between PM_2.5_, the source categories and ASCVD in exposure levels lower than the new annual NAAQS for PM_2.5_. First, we repeated the PM_2.5_ and ASVCD model. Then, we repeated the multivariable regression evaluating the associations of the five source categories with ASCVD. In this analysis, we report the associations per one unit increase to allow for direct comparisons amongst datasets.

All computational analyses were completed with the support of the Rollins High-Performance Computing (HPC) Cluster at Emory University and conducted using R (version 4.0.2) with the *mgcv*, *qgcomp*, and *meta* R packages.

## 3. Results

### 3.1. Descriptive Statistics

We included a total of 65,838,403 subjects with 559,591,306 person-years, 7.2% of whom had died of ASCVD, with larger mortality rates in the Northeast and Midwest. The mean age at entry was 65 years, about 56% were females, 84.7% were White, and 10.4% had Medicaid insurance (**Table 1**). **Table 2** shows the national and regional summary statistics of the exposures to PM_2.5_, its 15 chemical components, and the NMF source categories. Overall, the average PM_2.5_ mass level was 9.92 µg/m^3^. PM_2.5_ exposure was higher in the Midwest and Southeast.

**Table 1.**
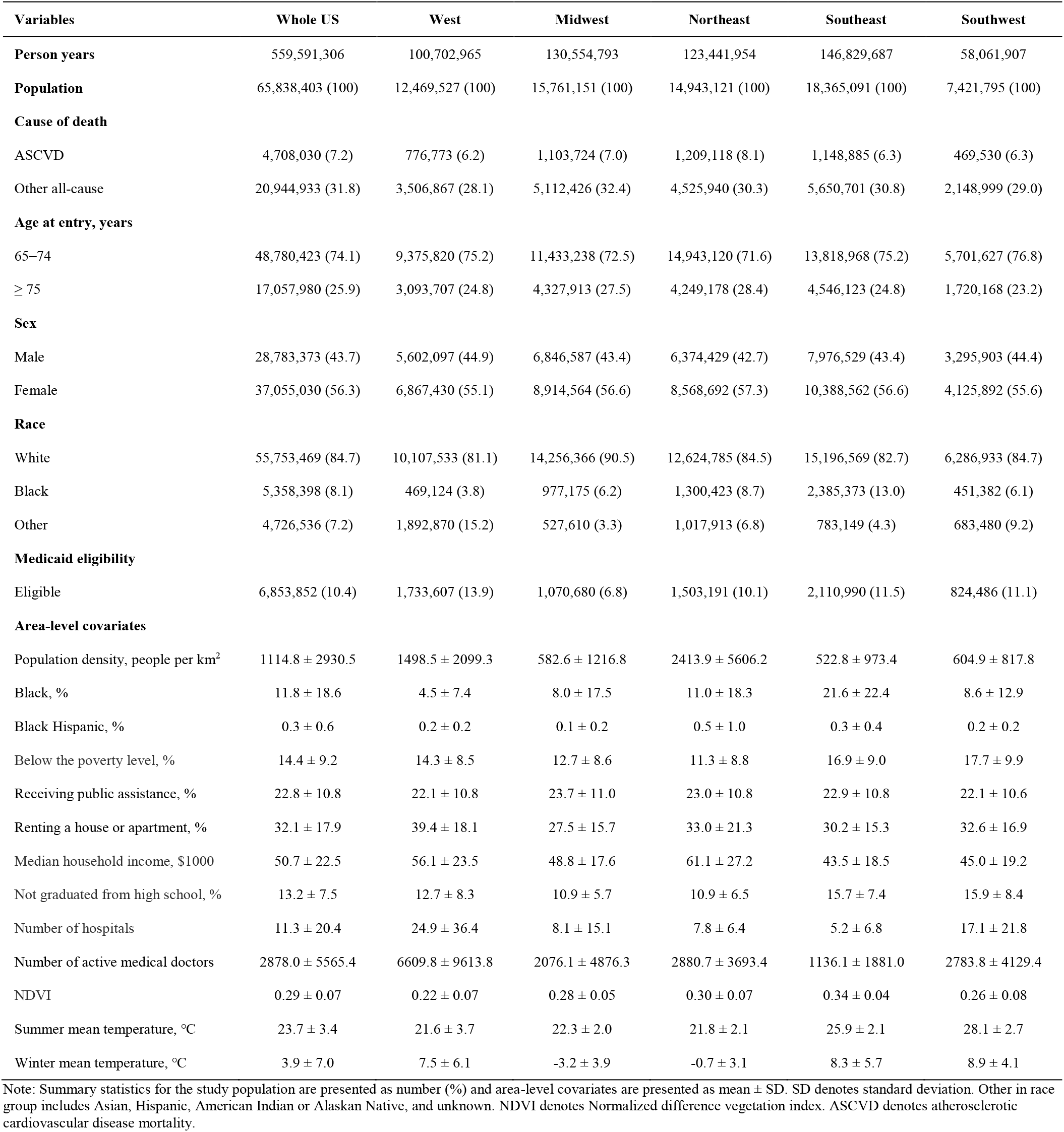
Descriptive statistics for the study population and area-level covariates among the cohort, by US region, and overall.

**Table 2.** Summary statistics of PM_2.5_ particle component concentrations and NMF loading factors over the study period, by US region and overall.

| Variable | Whole US. | West | Midwest | Northeast | Southeast | Southwest |
| --- | --- | --- | --- | --- | --- | --- |
| **PM<sub>2.5</sub> Major components** |  |  |  |  |  |  |
| SO<sub>4</sub><sup>2-</sup> (µg/m<sup>3</sup>) | 2.28 ± 1.17<br>(1.84) | 1.09 ± 0.66<br>(0.66) | 2.34 ± 1.00<br>(1.42) | 2.58 ± 1.10<br>(1.81) | 2.87 ± 1.10<br>(1.98) | 2.01 ± 0.93<br>(1.58) |
| NO<sub>3</sub><sup>-</sup> (µg/m<sup>3</sup>) | 1.09 ± 0.64<br>(0.89) | 1.34 ± 0.87<br>(1.23) | 1.61 ± 0.55<br>(0.81) | 1.10 ± 0.45<br>(0.62) | 0.67 ± 0.27<br>(0.32) | 0.72 ± 0.27<br>(0.35) |
| NH<sub>4</sub><sup>+</sup> (µg/m<sup>3</sup>) | 0.91 ± 0.48<br>(0.73) | 0.61 ± 0.48<br>(0.56) | 1.12 ± 0.44<br>(0.70) | 1.05 ± 0.49<br>(0.77) | 0.91 ± 0.43<br>(0.68) | 0.71 ± 0.31<br>(0.49) |
| OC (µg/m<sup>3</sup>) | 1.85 ± 0.68<br>(0.81) | 2.29 ± 1.04<br>(1.47) | 1.49 ± 0.37<br>(0.46) | 1.67 ± 0.44<br>(0.55) | 2.01 ± 0.53<br>(0.72) | 1.85 ± 0.58<br>(0.77) |
| EC (µg/m<sup>3</sup>) | 0.54 ± 0.27<br>(0.33) | 0.64 ± 0.37<br>(0.53) | 0.46 ± 0.20<br>(0.27) | 0.61 ± 0.30<br>(0.37) | 0.51 ± 0.18<br>(0.24) | 0.46 ± 0.24<br>(0.30) |
| **PM<sub>2.5</sub> Trace Elements** |  |  |  |  |  |  |
| Zn (ng/m<sup>3</sup>) | 7.82 ± 4.21<br>(4.78) | 7.12 ± 3.93<br>(5.12) | 9.04 ± 4.73<br>(5.27) | 9.98 ± 4.72<br>(5.23) | 6.17 ± 2.55<br>(2.93) | 6.62 ± 3.29<br>(4.22) |
| V (ng/m<sup>3</sup>) | 0.98 ± 0.89<br>(0.92) | 0.95 ± 0.85<br>(1.07) | 0.57 ± 0.41<br>(0.47) | 1.34 ± 1.22<br>(1.57) | 1.08 ± 0.85<br>(0.99) | 0.91 ± 0.67<br>(0.72) |
| K (ng/m<sup>3</sup>) | 58.89 ± 14.75<br>(17.26) | 60.58 ± 17.17<br>(17.29) | 53.69 ± 9.40<br>(11.00) | 48.63 ± 7.63<br>(8.71) | 63.64 ± 10.90<br>(13.85) | 72.96 ± 18.97<br>(19.26) |
| Si (ng/m<sup>3</sup>) | 104.77 ± 55.69<br>(69.85) | 116.40 ± 59.04<br>(72.74) | 85.49 ± 33.70<br>(41.58) | 60.65 ± 15.93<br>(20.98) | 107.70 ± 29.52<br>(46.81) | 193.59 ± 63.95<br>(57.15) |
| Pb (ng/m<sup>3</sup>) | 2.21 ± 1.27<br>(1.59) | 1.94 ± 1.16<br>(1.69) | 2.63 ± 1.48<br>(2.01) | 2.72 ± 1.42<br>(2.18) | 1.86 ± 0.90<br>(1.05) | 1.76 ± 0.87<br>(0.99) |
| Ni (ng/m<sup>3</sup>) | 0.63 ± 0.64<br>(0.55) | 0.65 ± 0.52<br>(0.79) | 0.40 ± 0.31<br>(0.49) | 1.10 ± 1.06<br>(0.95) | 0.53 ± 0.32<br>(0.41) | 0.43 ± 0.32<br>(0.42) |
| Fe (ng/m<sup>3</sup>) | 61.70 ± 28.98<br>(34.52) | 76.00 ± 36.82<br>(55.78) | 53.54 ± 24.22<br>(28.18) | 55.40 ± 26.84<br>(38.83) | 56.06 ± 17.81<br>(23.83) | 79.36 ± 32.65<br>(36.05) |
| Cu (ng/m<sup>3</sup>) | 2.46 ± 1.74<br>(2.38) | 3.36 ± 2.28<br>(3.84) | 1.83 ± 1.30<br>(1.80) | 2.64 ± 1.54<br>(2.17) | 2.18 ± 1.35<br>(1.86) | 2.60 ± 1.97<br>(2.74) |
| Ca (ng/m<sup>3</sup>) | 46.05 ± 25.93<br>(28.12) | 51.85 ± 30.92<br>(31.06) | 48.39 ± 18.52<br>(25.12) | 27.96 ± 9.24<br>(11.57) | 38.34 ± 12.88<br>(16.67) | 82.71 ± 30.64<br>(34.78) |
| Br (ng/m<sup>3</sup>) | 2.76 ± 0.68<br>(0.81) | 2.67 ± 1.03<br>(1.53) | 2.64 ± 0.68<br>(0.92) | 2.82 ± 0.58<br>(0.81) | 2.91 ± 0.41<br>(0.56) | 2.68 ± 0.56<br>(0.68) |
| **Total PM<sub>2.5</sub> Mass** |  |  |  |  |  |  |
| PM<sub>2.5</sub> (µg/m<sup>3</sup>) | 9.92 ± 3.17<br>(4.07) | 8.91 ± 4.48<br>(5.63) | 10.50 ± 2.77<br>(3.70) | 9.94 ± 2.84<br>(4.16) | 10.41 ± 2.58<br>(3.91) | 9.21 ± 2.70<br>(3.33) |
| **NMF Loadings** |  |  |  |  |  |  |
| Oil combustion | 7.44 ± 4.15<br>(4.03) | 8.17 ± 3.56<br>(4.20) | 7.48 ± 2.72<br>(3.23) | 10.31 ± 5.91<br>(5.32) | 5.27 ± 2.70<br>(3.34) | 6.25 ± 2.42<br>(3.22) |
| Soil/dust | 7.53 ± 4.23<br>(5.44) | 7.94 ± 4.34<br>(4.89) | 6.60 ± 2.91<br>(4.11) | 3.67 ± 1.39<br>(1.48) | 8.13 ± 2.31<br>(3.41) | 13.99 ± 4.55<br>(4.54) |
| Industrial | 6.13 ± 3.06<br>(4.10) | 3.72 ± 2.15<br>(2.10) | 9.33 ± 2.70<br>(3.50) | 6.84 ± 2.37<br>(2.86) | 5.15 ± 2.28<br>(3.09) | 4.92 ± 2.14<br>(2.95) |
| Coal/Biomass<br>burning | 9.74 ± 5.41<br>(7.55) | 8.41 ± 4.17<br>(6.25) | 5.20 ± 3.27<br>(4.55) | 11.01 ± 4.96<br>(7.07) | 13.65 ± 4.81<br>(6.69) | 8.83 ± 4.94<br>(6.93) |
| Motor vehicle | 9.26 ± 5.93<br>(7.42) | 14.59 ± 7.44<br>(11.65) | 10.12 ± 4.82<br>(6.54) | 8.09 ± 4.83<br>(6.24) | 6.66 ± 3.83<br>(4.57) | 7.77 ± 5.51<br>(7.10) |

Note: Summary statistics for air pollution concentrations are presented as mean ± SD (IQR). SD denotes standard deviation; IQR denotes interquartile range.

### 3.2. Non-negative matrix factorization (NMF) of 15 PM_2.5_ components

Based on the NMF results we labeled the 1^st^ category as Oil Combustion based on high loadings of Ni and V, the 2^nd^ category as Soil and Dust based on high loadings of Si and Ca, the 3^rd^ category as Industrial Pollution based on high loadings of SO_4_^2-^, NO_3_^-^, Z, and Pb, the 4^th^ category as Coal and Biomass Burning based on high loadings of OC and Br, and the 5^th^ category as Motor Vehicle based on high loadings of EC, Cu, and Fe (**Supplementary** Figure 2). Exposure levels of the different source categories also differed by region, with higher exposure to Oil Combustion in the Northeast, higher exposure to Soil and Dust in the Southwest, higher exposure to Industrial Pollution in the Midwest, higher exposure to Coal and Biomass Burning in the Southeast, and higher exposure to Motor Vehicle in the West (**Table 2**).

The correlation among the 15 PM_2.5_ components varied greatly, with the highest correlation between Cu and Fe (r=0.82), Cu and EC (r=0.78), Fe and EC (r=0.76), Si and Ca (r=0.79), V and Ni (r=0.73), SO_4_^2-^ and NH_4_ (r=0.84), and Z and Pb (r=0.75) (**Supplementary** Figure 3). The correlation between the source categories was low to moderate, with the highest correlation between the Oil Combustion and Soil and Dust sources (r=-0.39) (**Supplementary** Figure 4).

Higher levels of exposure to the Oil Combustion source category were associated with increased ASCVD mortality risk across most of the US, except for the West. Similarly, high levels of industrial pollution were associated with most US states except for the Northeast. PM_2.5_ attributed to Soil and Dust sources was associated with higher ASCVD risk only in the eastern regions and the Midwest. ASCVD mortality associations with Coal and Biomass burning PM_2.5_ were present in the western regions. Finally, ASCVD mortality associations with the Motor Vehicle source category were similar across all US regions, except for the Southeast. Comparing effect sizes of the source categories within each region: oil combustion was associated with the largest ASCVD mortality risk in the Northeast (RR 1.094, 95% CI 1.088;01.100), Southeast (RR 1.089, 95% CI 1.085; 1.093), and Midwest (RR1.079, 95% CI 1.075; 1.084); and Coal and Biomass burning exposure was associated with the largest ASCVD mortality risk in the West (RR 1.086, 95% CI 1.079; 1.094) and Southwest (RR 1.071, 95% CI 1.062; 1.080) (**Figure 1 and Supplementary Table 1**).

**Figure 1.**
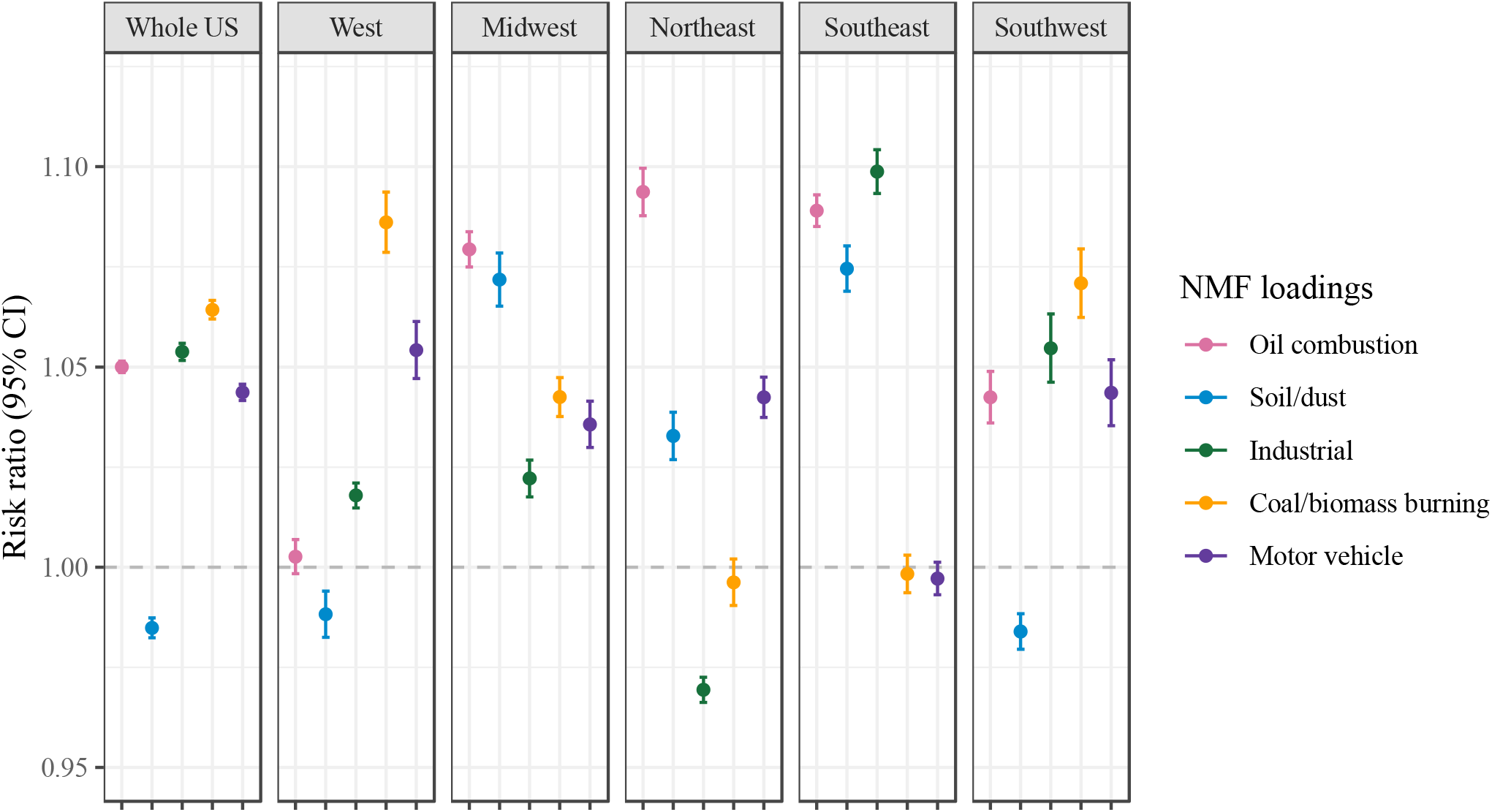
Risk ratio of ASCVD mortality per IQR increase in each NMF loading factor by US regions and overall. The estimated risk ratios were obtained using the multivariate Poisson regression model. Error bars represent the 95% confidence intervals. ASCVD denotes atherosclerotic cardiovascular disease; IQR denotes the interquartile range; NMF denotes non-negative matrix factorization.

### 3.3 Subset below 9 μg/m^3^ PM_2.5_

The subset includes 223,548,803 person-years in 36,879 ZIP codes. While most of the ZIP codes were included for at least a year in the subset, the Southeast and bordering areas were included in fewer years than the rest of the US (**Supplementary figure 5**). Concentrations of PM_2.5_ and most components were smaller in the subset, except for Si (**Supplementary Table 2**). Similarly, the means of the factors were smaller except for Soil and Dust (Si being a major component). Oil combustion, Soil and Dust, and Motor Vehicles associations are similar in both datasets, and Industrial and Coal and Biomass Burning associations were stronger in the bellow 9 μg/m^3^ PM_2.5_ dataset (**Supplementary** Figure 6; **Supplementary Table 4**).

## 4. Discussion

In this study, we aimed to delineate the associations between PM_2.5_ and its components and sources with ASCVD mortality, emphasizing individual and combined effects, as well as regional variations in the United States. We found higher ASCVD risk associated with oil combustion, coal or biomass burning, industrial, and motor vehicle sources.

The exposure patterns and associations varied regionally. Overall, PM_2.5_ mass exposure was highest in the Midwest and Southeast. Motor Vehicle sources dominated in the Midwest, and coal and biomass-burning sources dominated in the Northeast. In Midwest and Northeast associations with oil combustion sources were more pronounced compared to other sources. In the Southeast, coal and biomass-burning was a dominant source of exposure, and the associations with industrial sources were more pronounced than with other sources. In the West, traffic pollution was dominant. In the sources analysis, the association with coal and biomass burning sources was more pronounced compared to other sources. Finally, in the Southwest, soil and dust sources were dominant. The association with coal and biomass burning sources was more pronounced compared to other sources.

### PM_2.5_ Sources and ASCVD mortality

Our NMF results indicated that oil, industrial, coal, biomass burning, and traffic sources were associated with increased ASCVD mortality risk across the US. Soil and Dust was not associated with higher ASCVD mortality risk. It is challenging to draw direct comparisons with other studies that use factorization methods to group components into sources because different studies use different factors. Yet, multiple studies find adverse cardiovascular outcomes associated with oil combustion, industrial, biomass, coal burning, and motor vehicle sources. In a 2022 study, Kazemiparkouhi et al. categorized 18 components into eight factors and found that cardiovascular mortality had the strongest associations with traffic and coal. However, unlike our study, they also found strong associations with soil sources ^14^. Like our study, Henneman et al. highlight the mortality risk associated with coal-sourced PM_2.5_ among the Medicare population in the United States ^24^. Moreover, Thurston et al. found substantial associations between coal combustion and traffic-related sources and ischemic heart disease. However, they did not find associations with soil and biomass combustion ^25^. Lastly, Hill et al. found associations between industrial pollution and children’s cardiovascular functioning ^26^.

Our results consistently pointed toward an effect modification by US region in the sources-centered analysis. In the Northeast and Midwest, the associations with oil combustion sources were more pronounced compared to other sources. In the West and Southwest, the associations with coal and biomass burning were more pronounced compared to other sources. These regional differences require further investigation as these can be attributed to a combination of diverse factors. For example, each US region has a distinct climate with summer mean temperature varying from 21.6 °C in the West to 28.1 °C in the Southwest. Temperature ^27^, as well as other climate variables ^28^, were found to modify PM_2.5_-associated health effects. Furthermore, components of the built environment can also influence the association. Greenness levels, which were found to attenuate the effects of PM_2.5_ ^29^, are lower on average in the West compared to the Southeast. Additionally, social and demographic characteristics of the population and differential prevalence of vulnerable populations of racial/ethnic minorities and low socioeconomic status, can contribute to regional differences ^30,31^. Finally, the uneven distribution of ASCVD incidence and its underling risk factors should also be considered. For example, the Southeastern region in the US is known as the “Stroke Belt” due to significant clustering of stroke incident cases in these states. This heterogeneous incidence distribution can be linked to differences in population vulnerability, socioeconomic status, race and ethnicity as well as environmental and behavioral factors ^32^.

### Regional variations and socioeconomic implications

The regional disparities in PM_2.5_ exposure and its association with ASCVD mortality underscore the intricate interplay between environmental factors and social determinants of health. Populations in the West and Midwest, for instance, showed larger associations with ASCVD, which might reflect regional differences in industrial activity, urbanization, and environmental regulations. This adds to the context of marginalized communities, including racial/ethnic minorities ^33^ and individuals with lower socioeconomic status ^34^, being disproportionally impacted by air pollution, either by higher exposure or reduced access to care. These inequalities are more present in some regions than others ^15^, which could be a consequence of regional differences in customs, traditions, and lifestyle that can also influence this association and are challenging to capture in retrospective epidemiological studies.

### Implications for Public Health and Policy

Our findings have significant implications for public health and policy-making. The clear link between PM_2.5_ exposure and ASCVD mortality warrants enhanced efforts in air quality improvement. Identifying specific PM_2.5_ sources as major contributors to ASCVD mortality offers a targeted approach for environmental regulations. The observed disparities in pollution exposure and health impacts call for more equitable environmental and health policies that integrate localized data, prioritize health outcomes, and address the needs of the most vulnerable populations. All these implications should also be considered within each region-specific context, as our results highlight the differences in the associations between regions.

Additionally, our results centering on ZIP-year below the new NAAQS of 9 μg/m^3^ PM_2.5_ showed that PM_2.5_ is still detrimental to health even when <9 μg/m^3^. These results are consistent with previous studies that found associations between PM_2.5_ at <12 μg/m^3^ and excess mortality ^35^, as well as excess CVD mortality in low-exposure regions of North Carolina^36^ and Canada^37^. These results highlight the need to close the gap between the NAAQS and the WHO recommendation – currently set at <5 μg/m^3^.

### Strengths and limitations

In this study, we conducted a national analysis including diverse older adults in the Medicare populations and exposure characteristics. We used tailored statistical methods for mixtures capable of handling the high complexity and correlation structure of the components data. We leveraged high-resolution geospatial models of numerous PM_2.5_ components. This adds crucial evidence to current research largely focused on a limited set of PM_2.5_ components due to restricted data availability^38^ or limited geographic areas^39^.

Our study is not without limitations. Firstly, despite the high performance of our exposure assessment models, the reliance on modeled pollutant concentrations introduces some measurement errors, and the use of ZIP code level exposure might introduce some exposure misclassification. Second, while our statistical models accounted for many potential confounders, the lack of certain individual-level risk factors, such as smoking, drug use, and alcohol consumption, which are linked to ASCVD mortality, might have influenced our risk estimates. However, since the exposure is assigned on a ZIP code level we do not expect this to bias our results.

## 5. Conclusions

Our study provides a comprehensive assessment of the associations between PM_2.5_ chemical components sources and ASCVD mortality, highlighting both individual and combined effects, as well as distinct regional variations across the United States. We established a link between PM_2.5_ and ASCVD mortality, identifying specific components and sources as key drivers of this association. Furthermore, our findings showcase regional heterogeneity, with stronger associations in certain areas. These associations persisted even after limiting our sample to ZIP code-years with PM_2.5_ <9 μg/m^3^ – the new NAAQS. These results emphasize the role of geographical context and the need to consider local population characteristics and exposure patterns when assessing health risks associated with air pollution.

## Data Availability

Restrictions apply to the availability of these data, which were used under license for this study.

## Acknowledgements

This study was supported by the HERCULES Center (P30 ES019776), the Mount Sinai transdisciplinary center on early environmental exposures (P30 ES023515 and P30 AG021342), the National Institute on Aging (NIA/NIH R01 AG074357), the National Institute of Environmental Health Sciences (R21 ES032606, R01 ES032242, 5U2CES026555-03, R01 ES013744, P30 ES000002, R01 ES032418, and UL1TR004419), and the United States Environmental Protection Agency (US EPA) (RD-83587201). Its contents are solely the responsibility of the grantee and do not necessarily represent the official views of the US EPA. Furthermore, the US EPA does not endorse the purchase of any commercial products or services mentioned in the publication.

## References

1. Van Trier T, Mohammadnia N, Snaterse M, Peters R, Jørstad H, Bax W. Lifestyle management to prevent atherosclerotic cardiovascular disease: evidence and challenges. Netherlands Heart Journal 2022;30(1):3–14.

2. Münzel T, Hahad O, Sørensen M, et al. Environmental risk factors and cardiovascular diseases: a comprehensive expert review. Cardiovascular Research 2022;118(14):2880–2902.

3. Krittanawong C, Qadeer YK, Hayes RB, et al. PM2. 5 and cardiovascular health risks. Current problems in cardiology 2023;48(6):101670.

4. Alexeeff SE, Liao NS, Liu X, Van Den Eeden SK, Sidney S. Long-term PM2. 5 exposure and risks of ischemic heart disease and stroke events: review and meta-analysis. Journal of the American Heart Association 2021;10(1):e016890.

5. Abubakar I, Tillmann T, Banerjee A. Global, regional, and national age-sex specific all-cause and cause-specific mortality for 240 causes of death, 1990-2013: a systematic analysis for the Global Burden of Disease Study 2013. Lancet 2015;385(9963):117-171.

6. Liu Y-H, Bo Y-C, You J, Liu S-F, Liu M-J, Zhu Y-J. Spatiotemporal trends of cardiovascular disease burden attributable to ambient PM2. 5 from 1990 to 2019: A global burden of disease study. Science of The Total Environment 2023;885:163869.

7. Li B, Ma Y, Zhou Y, Chai E. Research progress of different components of PM2. 5 and ischemic stroke. Scientific Reports 2023;13(1):15965.

8. Basagaña X, Jacquemin B, Karanasiou A, et al. Short-term effects of particulate matter constituents on daily hospitalizations and mortality in five South-European cities: Results from the MED-PARTICLES project. Environment International 2015;75:151–158.

9. Ferreira TM, Forti MC, De Freitas CU, Nascimento FP, Junger WL, Gouveia N. Effects of particulate matter and its chemical constituents on elderly hospital admissions due to circulatory and respiratory diseases. International journal of environmental research and public health 2016;13(10):947.

10. Lin H, Tao J, Qian ZM, et al. Shipping pollution emission associated with increased cardiovascular mortality: A time series study in Guangzhou, China. Environmental Pollution 2018;241:862–868.

11. Jin T, Amini H, Kosheleva A, et al. Associations between long-term exposures to airborne PM2. 5 components and mortality in Massachusetts: mixture analysis exploration. Environmental Health 2022;21(1):1-13.

12. Yazdi MD, Amini H, Wei Y, Castro E, Shi L, Schwartz JD. Long-term exposure to PM2. 5 species and all-cause mortality among Medicare patients using mixtures analyses. Environmental Research 2024;246:118175.

13. Yang Q, Cogswell ME, Flanders WD, et al. Trends in cardiovascular health metrics and associations with all-cause and CVD mortality among US adults. Jama 2012;307(12):1273–1283.

14. Kazemiparkouhi F, Honda T, Eum K-D, Wang B, Manjourides J, Suh HH. The impact of Long-Term PM2. 5 constituents and their sources on specific causes of death in a US Medicare cohort. Environment International 2022;159:106988.

15. Knobel P, Hwang I, Castro E, et al. Socioeconomic and racial disparities in source-apportioned PM2. 5 levels across urban areas in the contiguous US, 2010. Atmospheric Environment 2023;303:119753.

16. Tibuakuu M, Michos ED, Navas-Acien A, Jones MR. Air pollution and cardiovascular disease: a focus on vulnerable populations worldwide. Current epidemiology reports 2018;5:370–378.

17. Amini H, Danesh-Yazdi M, Di Q, et al. Hyperlocal super-learned PM2. 5 components across the contiguous US. 2022.

18. Amini H, Danesh-Yazdi M, Di Q, et al. Hyperlocal US PM2. 5 Trace Elements Super-learned. 2022.

19. Amini H, Danesh-Yazdi M, Di Q, Requia WJ, Wei Y, AbuAwad Y. Annual Mean PM2.5 Trace Elements 50m Grids in Urban Areas and 1km Grids in Non-Urban Areas for Contiguous U.S., 2000-2019, v1. (Preliminary Release). Palisades, New York: NASA Socioeconomic Data and Applications Center (SEDAC); 2022.

20. Amini H, Danesh-Yazdi M, Di Q, Requia WJ, Wei Y, AbuAwad Y. Annual Mean PM2.5 Components (EC, NH4, NO3, OC, SO4) 50m Urban and 1km Non-Urban Area Grids for Contiguous U.S., 2000-2019 v1. (Preliminary Release). Palisades, New York: NASA Socioeconomic Data and Applications Center (SEDAC); 2022.

21. Ma T, Yazdi MD, Schwartz J, et al. Long-term air pollution exposure and incident stroke in American older adults: A national cohort study. Global Epidemiology 2022;4:100073.

22. Jin T, Amini H, Kosheleva A, et al. Associations between long-term exposures to airborne PM(2.5) components and mortality in Massachusetts: mixture analysis exploration. Environ Health 2022;21(1):96. (In eng). DOI: 10.1186/s12940-022-00907-2.

23. Whitehead J. Fitting Cox’s regression model to survival data using GLIM. Journal of the Royal Statistical Society Series C: Applied Statistics 1980;29(3):268–275.

24. Henneman L, Choirat C, Dedoussi I, Dominici F, Roberts J, Zigler C. Mortality risk from United States coal electricity generation. Science 2023;382(6673):941-946.

25. Thurston GD, Burnett RT, Turner MC, et al. Ischemic heart disease mortality and long-term exposure to source-related components of US fine particle air pollution. Environmental health perspectives 2016;124(6):785–794.

26. Hill DT, Petroni M, Larsen DA, et al. Linking metal (Pb, Hg, Cd) industrial air pollution risk to blood metal levels and cardiovascular functioning and structure among children in Syracuse, NY. Environmental Research 2021;193:110557.

27. Li Y, Ma Z, Zheng C, Shang Y. Ambient temperature enhanced acute cardiovascular-respiratory mortality effects of PM 2.5 in Beijing, China. International journal of biometeorology 2015;59:1761–1770.

28. Ito K, Thurston GD, Silverman RA. Characterization of PM2. 5, gaseous pollutants, and meteorological interactions in the context of time-series health effects models. Journal of exposure science & environmental epidemiology 2007;17(2):S45-S60.

29. Yitshak-Sade M, James P, Kloog I, et al. Neighborhood greenness attenuates the adverse effect of PM2. 5 on cardiovascular mortality in neighborhoods of lower socioeconomic status. International journal of environmental research and public health 2019;16(5):814.

30. Han C, Xu R, Gao CX, et al. Socioeconomic disparity in the association between long-term exposure to PM2. 5 and mortality in 2640 Chinese counties. Environment international 2021;146:106241.

31. Ma Y, Zang E, Opara I, Lu Y, Krumholz HM, Chen K. Racial/ethnic disparities in PM2. 5-attributable cardiovascular mortality burden in the United States. Nature Human Behaviour 2023:1-10.

32. Howard G, Howard VJ. Twenty years of progress toward understanding the stroke belt. Stroke 2020;51(3):742–750.

33. Spiller E, Proville J, Roy A, Muller NZ. Mortality risk from PM 2.5: a comparison of modeling approaches to identify disparities across racial/ethnic groups in policy outcomes. Environmental health perspectives 2021;129(12):127004.

34. Hajat A, Hsia C, O’Neill MS. Socioeconomic disparities and air pollution exposure: a global review. Current environmental health reports 2015;2:440–450.

35. Di Q, Wang Y, Zanobetti A, et al. Air pollution and mortality in the Medicare population. New England Journal of Medicine 2017;376(26):2513–2522.

36. Weichenthal S, Villeneuve PJ, Burnett RT, et al. Long-term exposure to fine particulate matter: association with nonaccidental and cardiovascular mortality in the agricultural health study cohort. Environmental health perspectives 2014;122(6):609–615.

37. Crouse DL, Peters PA, van Donkelaar A, et al. Risk of nonaccidental and cardiovascular mortality in relation to long-term exposure to low concentrations of fine particulate matter: a Canadian national-level cohort study. Environmental health perspectives 2012;120(5):708–714.

38. Yang Y, Ruan Z, Wang X, et al. Short-term and long-term exposures to fine particulate matter constituents and health: a systematic review and meta-analysis. Environmental pollution 2019;247:874–882.

39. Karimi B, Samadi S. Mortality associated with fine particulate and its components: A systematic review and meta-analysis. Atmospheric Pollution Research 2023:101648.

