## Supplemental Files for "PM_2.5_ components mixture and atherosclerotic cardiovascular disease mortality: a national analysis of Medicare enrollees"

**Supplementary Figure 1.** Map of 5 geographical regions (West, Midwest, Northeast, Southeast, Southwest) in the contiguous US, and summary of results.

**West**

Exposure patterns:

- Highest motor vehicle contribution.

Associations with sources:

- Higher risks associated with coal/biomass, motor vehicle, industrial and oil sources.
- Largest association with coal/biomass burning sources.

**Midwest**

Exposure patterns:

- Higher $PM_{2.5}$ mass exposure overall.
- Highest industrial source contribution.

Associations with sources:

- Higher risks associated with all the five sources.
- Largest association with oil sources.

**Northeast**

Exposure patterns:

- Highest oil source contribution.

Associations with sources:

- Higher risks associated with oil, motor vehicle, and soil/dust sources.
- Largest associations with oil sources.

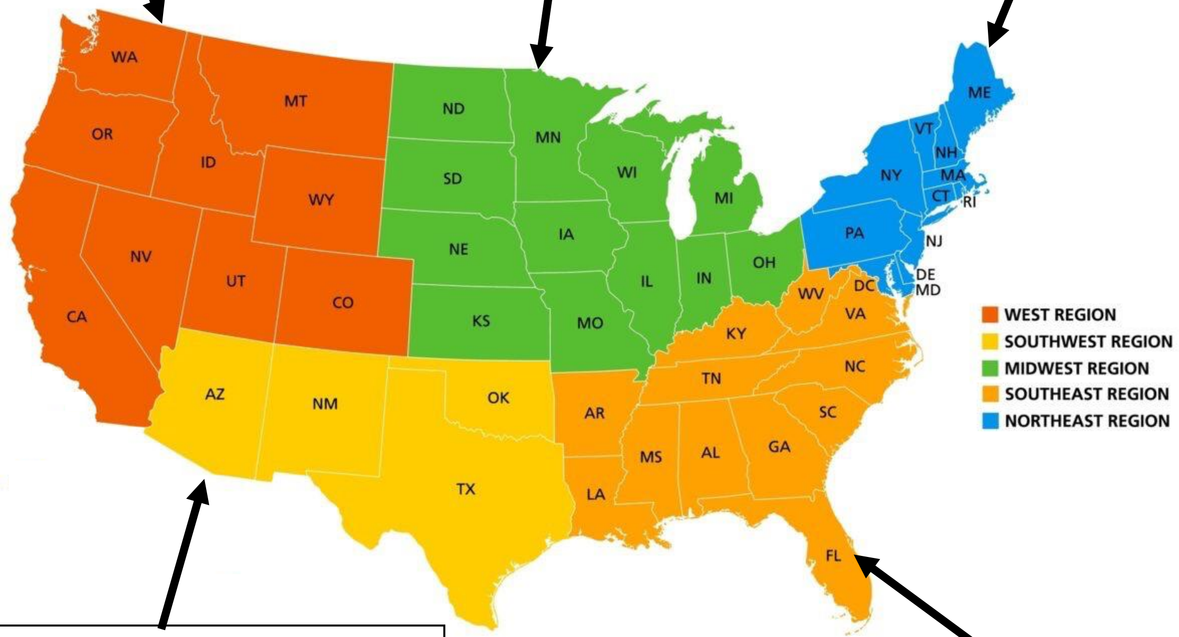


**Southwest**

Exposure patterns:

- Highest soil/dust source contribution.

Associations with sources:

- Higher risk associated with coal/biomass, industrial, motor vehicle, and oil sources.
- Largest associations with coal/biomass sources.

**Southeast**

Exposure patterns:

- Higher $PM_{2.5}$ mass exposure overall.
- Highest coal/biomass source contribution.

Associations with sources:

- Higher risk associated with industrial, soil/dust, and oil sources.
- Largest association with oil sources.

**Supplementary Figure 2.** Heat map of the mixture coefficient matrix.

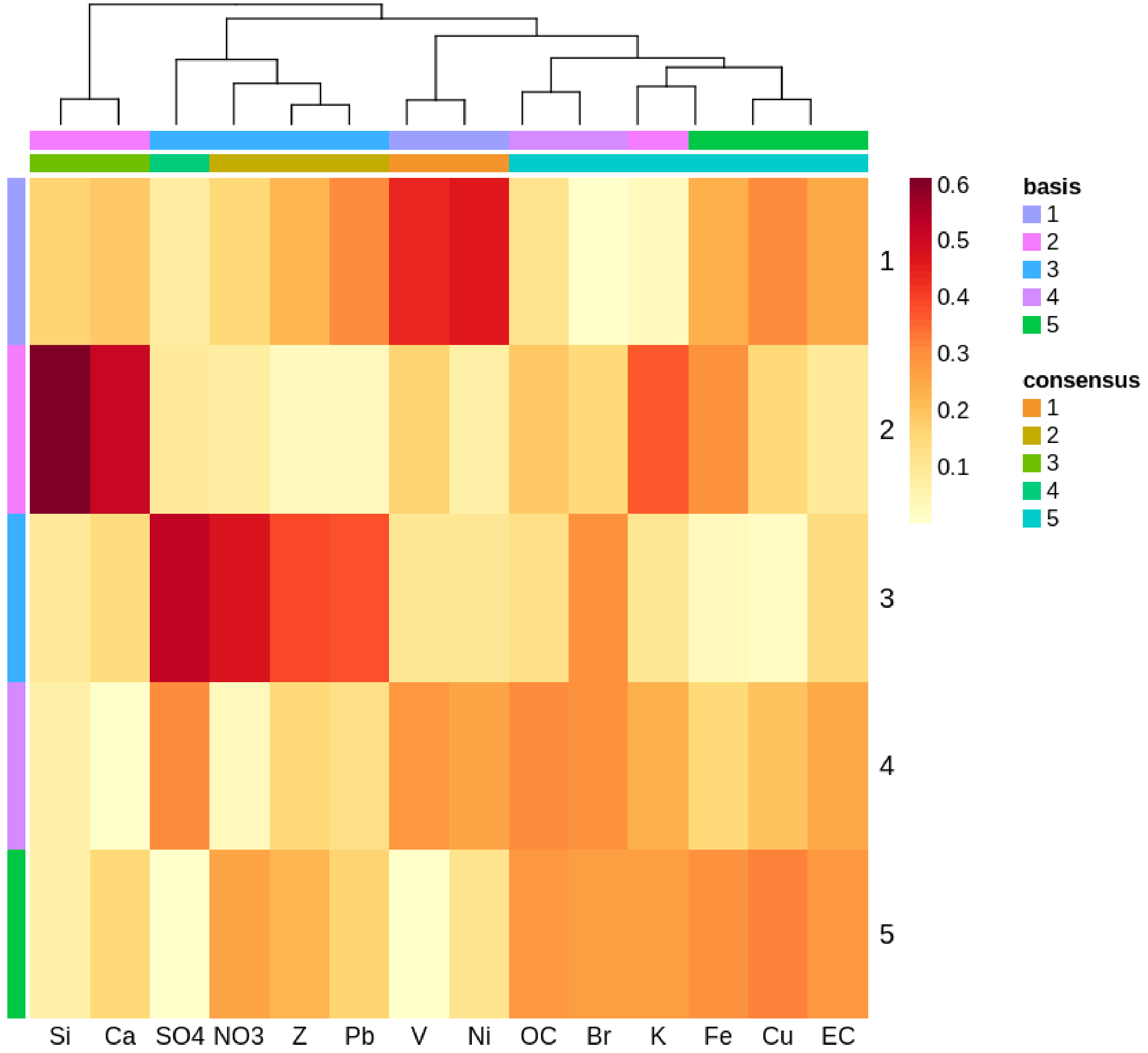


Each column corresponds to a $PM_{2.5}$ component. We label the 1st category as Oil Combustion based on high loadings of nickel (Ni) and vanadium (V), the 2nd category as Soil and Dust based on high loadings of silicon (Si) and calcium (Ca), the 3rd category as Industrial Pollution based on high loadings of sulfate ($SO_4^{2-}$), nitrate ($NO_3^-$), zinc (Z), and lead (Pb), the 4th category as Coal and Biomass Burning based on high loadings of organic carbon (OC) and bromine (Br), and the 5th category as Motor Vehicle based on high loadings of elemental carbon (EC), cupper (Cu), and iron (Fe).

**Supplementary Figure 3.** Correlation matrix among the 15 $PM_{2.5}$ components.

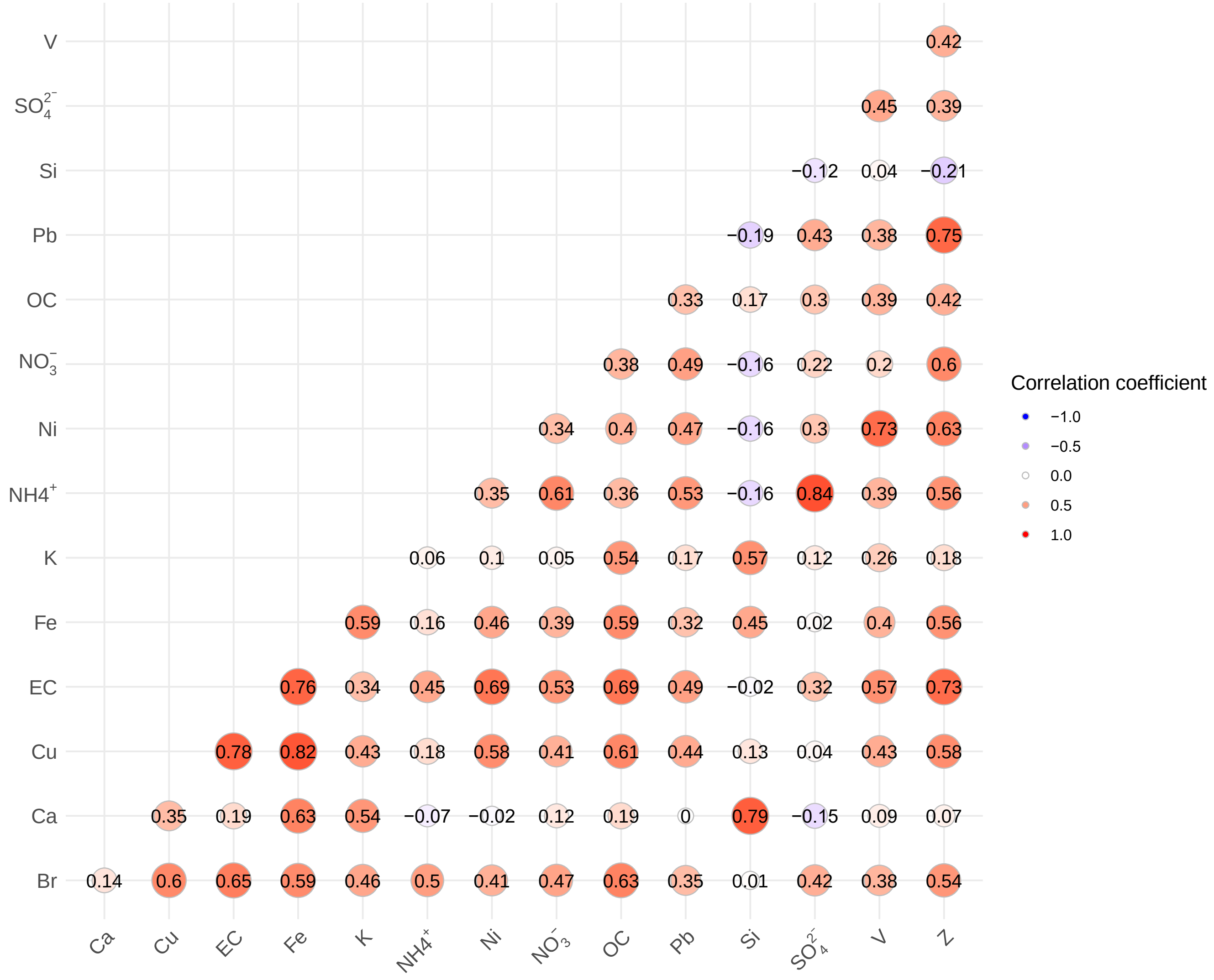

**Supplementary Figure 4.** Correlation matrix among the 5 NMF loading factors.

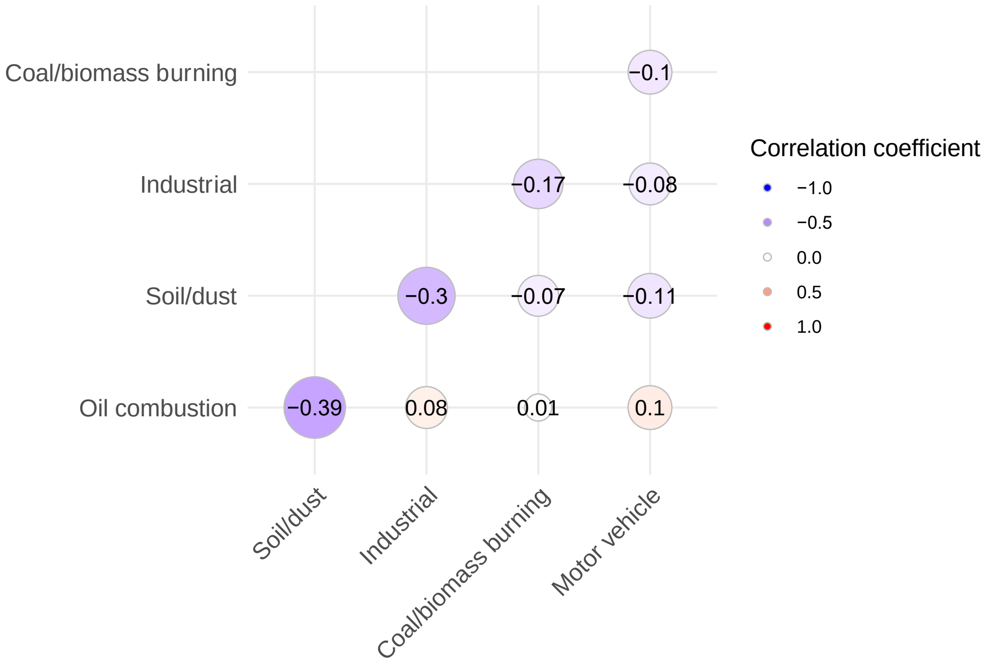


**Supplementary Table 1.** Risk ratio of cardiovascular mortality per interquartile range (IQR) increase in each non-negative matrix factorization (NMF) loading factor from multi-variate Cox regression models by US regions and overall. The effect estimates are presented in Figure 1.

| NMF loadings | Whole US | West | Midwest | Northeast | Southeast | Southwest |
|---|---|---|---|---|---|---|
| Oil combustion | 1.050 (1.049, 1.051) | 1.003 (0.998, 1.007) | 1.079 (1.075, 1.084) | 1.094 (1.088, 1.100) | 1.089 (1.085, 1.093) | 1.042 (1.036, 1.049) |
| Soil/dust | 0.985 (0.982, 0.987) | 0.998 (0.982, 0.994) | 1.072 (1.065, 1.078) | 1.033 (1.027, 1.039) | 1.075 (1.069, 1.080) | 0.984 (0.980, 0.988) |
| Industrial | 1.054 (1.052, 1.056) | 1.018 (1.015, 1.021) | 1.022 (1.018, 1.027) | 0.969 (0.966, 0.973) | 1.099 (1.093, 1.104) | 1.055 (1.046, 1.063) |
| Coal/biomass burning | 1.064 (1.062, 1.067) | 1.086 (1.079, 1.094) | 1.042 (1.038, 1.047) | 0.996 (0.990, 1.002) | 0.998 (0.994, 1.003) | 1.071 (1.062, 1.080) |
| Motor vehicle | 1.044 (1.042, 1.046) | 1.054 (1.047, 1.061) | 1.036 (1.030, 1.041) | 1.042 (1.037, 1.047) | 0.997 (0.993, 1.001) | 1.044 (1.035, 1.052) |

**Supplementary Figure 5:** Map A shows ZIPs included in low-exposure dataset shaded in blue. Map B shoes how many years of data were included for each ZIP.

A

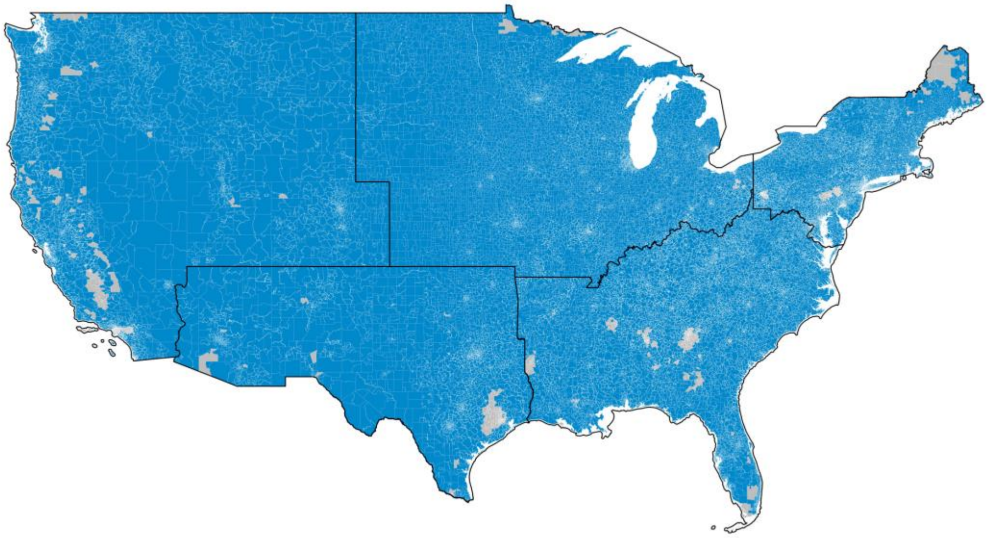

B

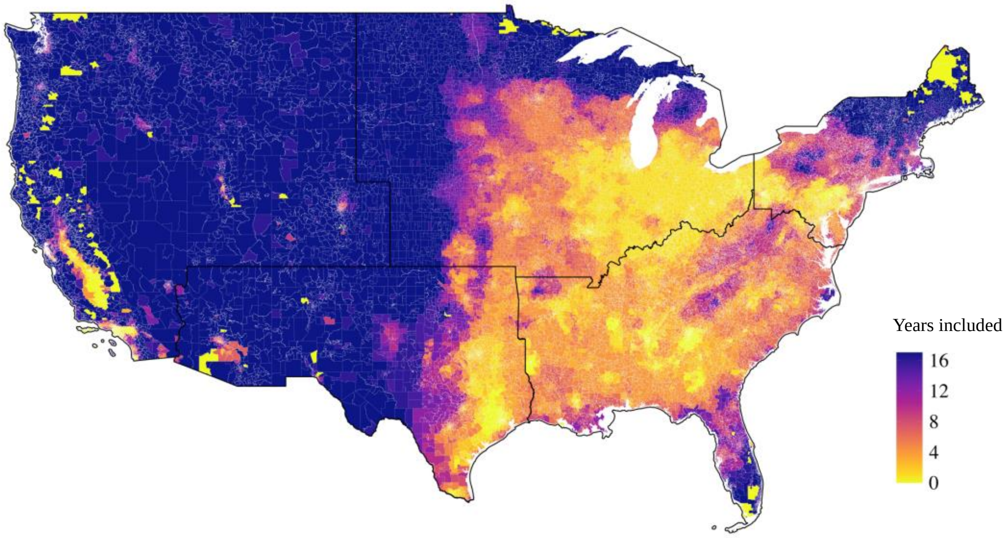

**Supplementary Table 2.** Summary statistics of $PM_{2.5}$ particle component concentrations and NMF loading factors over the study period comparing full dataset and subset when $PM_{2.5}$ is restricted to less than 9 μg/m$^3$.

| Dataset | Full range exposure | Low-exposure |
|---|---|---|
| **$PM_{2.5}$ Major Components** | | |
| $SO_4^{2-}$ (μg/m$^3$) | 2.28 ± 1.17 (1.84) | 1.37 ± 0.66 (0.90) |
| $NO_3^-$ (μg/m$^3$) | 1.09 ± 0.64 (0.89) | 0.75 ± 0.39 (0.50) |
| $NH_4^+$ (μg/m$^3$) | 0.91 ± 0.48 (0.73) | 0.51 ± 0.24 (0.34) |
| OC (μg/m$^3$) | 1.85 ± 0.68 (0.81) | 1.48 ± 0.49 (0.62) |
| EC (μg/m$^3$) | 0.54 ± 0.27 (0.33) | 0.40 ± 0.21 (0.27) |
| **$PM_{2.5}$ Trace Elements** | | |
| Zn (ng/m$^3$) | 7.82 ± 4.21 (4.78) | 5.55 ± 2.89 (3.34) |
| V (ng/m$^3$) | 0.98 ± 0.89 (0.92) | 0.65 ± 0.67 (0.51) |
| K (ng/m$^3$) | 58.89 ± 14.75 (17.26) | 53.63 ± 15.58 (18.29) |
| Si (ng/m$^3$) | 104.77 ± 55.69 (69.85) | 107.33 ± 66.59 (83.99) |
| Pb (ng/m$^3$) | 2.21 ± 1.27 (1.59) | 1.65 ± 1.12 (0.77) |
| Ni (ng/m$^3$) | 0.63 ± 0.64 (0.55) | 0.41 ± 0.38 (0.52) |
| Fe (ng/m$^3$) | 61.70 ± 28.98 (34.52) | 53.38 ± 27.44 (33.08) |
| Cu (ng/m$^3$) | 2.46 ± 1.74 (2.38) | 2.00 ± 1.63 (2.12) |
| Ca (ng/m$^3$) | 46.05 ± 25.93 (28.12) | 45.49 ± 30.08 (33.94) |
| Br (ng/m$^3$) | 2.76 ± 0.68 (0.81) | 2.35 ± 0.66 (0.90) |
| **Total $PM_{2.5}$ Mass** | | |
| $PM_{2.5}$ (μg/m$^3$) | 9.92 ± 3.17 (4.07) | 6.91 ± 1.66 (2.31) |
| **NMF Loadings** | | |
| Oil combustion | 7.44 ± 4.15 (4.03) | 7.27 ± 2.59 (2.79) |
| Soil/dust | 7.53 ± 4.23 (5.44) | 8.20 ± 4.79 (6.33) |
| Industrial | 6.13 ± 3.06 (4.10) | 4.61 ± 2.13 (2.91) |
| Coal/biomass burning | 9.74 ± 5.41 (7.55) | 7.35 ± 4.39 (6.23) |
| Motor vehicle | 9.26 ± 5.93 (7.42) | 8.54 ± 5.06 (6.44) |

Supplementary Table 3: Corresponding estimates for low-exposure dataset(2)

| **$PM_{2.5}$ mass** | **Whole US** | **West** | **Midwest** | **Northeast** | **Southeast** | **Southwest** |
|---|---|---|---|---|---|---|
| Full range | 1.012 (1.011, 1.012) | 1.013 (1.012, 1.014) | 1.007 (1.006, 1.009) | 0.985 (0.984, 0.987) | 0.964 (0.963, 0.966) | 1.010 (1.008, 1.013) |
| Below 9 μg/m$^3$ | 1.028 (1.026, 1.029) | 1.013 (1.011, 1.015) | 1.029 (1.024, 1.033) | 1.017 (1.013, 1.021) | 0.966 (0.962, 0.971) | 1.020 (1.015, 1.025) |

**Supplementary Figure 6.** Risk ratio of ASCVD mortality per one unit increase in each NMF loading factor in ZIP-years bellow 9 µg/m$^3$ $PM_{2.5}$. The estimated risk ratios were obtained using the multivariate Poisson regression model. Error bars represent the 95% confidence intervals. ASCVD denotes atherosclerotic cardiovascular disease; NMF denotes non-negative matrix factorization.

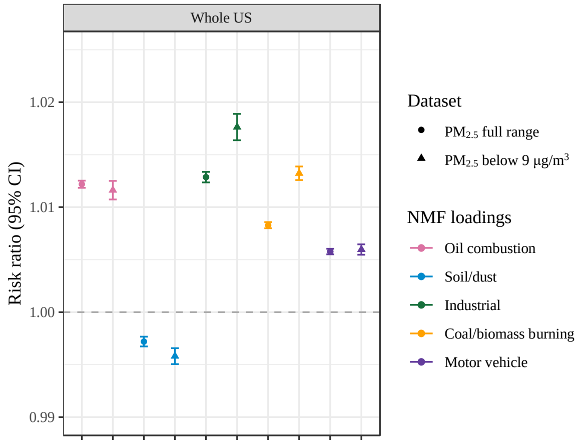

**Supplementary Table 4:** Risk ratio of ASCVD mortality per one unit increase in each NMF loading factor in ZIP-years bellow 9 μg/m$^3$ $PM_{2.5}$.. The estimated risk ratios were obtained using the multivariate Poisson regression model. Error bars represent the 95% confidence intervals. ASCVD denotes atherosclerotic cardiovascular disease; NMF denotes non-negative matrix factorization.

| NMF loadings | $PM_{2.5}$ full range dataset | $PM_{2.5}$ below 9 μg/m$^3$ |
|---|---|---|
| Oil combustion | 1.0122 (1.0118, 1.0125) | 1.0116 (1.0107, 1.0125) |
| Soil/dust | 0.9972 (0.9967, 0.9977) | 0.9958 (0.9951, 0.9966) |
| Industrial | 1.0129 (1.0124, 1.0134) | 1.0176 (1.0164, 1.0189) |
| Coal/biomass burning | 1.0083 (1.0080, 1.0086) | 1.0132 (1.0126, 1.0139) |
| Motor vehicle | 1.0058 (1.0055, 1.0060) | 1.0060 (1.0055, 1.0065) |
